# Leveraging Explainable Temporal-Modelling Machine Learning to Identify Distinct Multimorbidity Trajectory Profiles in Acute Myocardial Infarction

**DOI:** 10.64898/2026.01.14.26344136

**Authors:** Anthony Onoja, Kris Elomaa, Anthony D Whetton, Nophar Geifman

## Abstract

**Introduction:** Acute myocardial infarction (AMI) remains a leading cause of mortality, with the coexistence of other conditions (i.e., multimorbidity) complicating management and outcomes. Currently, healthcare providers see major challenges in consideration of the patient with a multimorbid profile, especially as this is a progressive issue where the temporal evolution of diseases is complex in nature, with a profound impact on clinical outcomes.

**Methods:** Data on 12,701 AMI patients from the UK Biobank were selected for analysis from the cohort of 502,000 volunteers and then grouped into pre- (up to 1 year prior) and early (within 5 years) post-AMI periods. Using Dynamic Time Warping (DTW) clustering, sequences of ICD-10 diagnoses accumulated over time in the post-AMI period were used to cluster participants. Topic modelling of cluster-specific diagnoses informed thematic labels for these profiles (clusters) of AMI patients. Using data from pre-AMI, along with socio-demographic variables (age, IMD score, BMI, and sex), four predictive supervised models, namely, Logistic Regression, Random Forest, XGBoost, and CatBoost, were developed, with CatBoost achieving the highest accuracy for profile membership prediction. Model interpretability via SHapley Additive exPlanations (SHAP) identified key diagnostic categories that were driving profile assignments. Then, survival analyses compared SMART (Second Manifestations of Arterial Disease) risk scores across the profiles, adjusting for clinical covariates to evaluate adverse cardiovascular outcomes - death. Finally, Phenome-Wide Association Studies (PheWAS) were employed to link profile-specific diagnostic themes to underlying genetic mechanisms.

**Results:** Using the above approaches, three multimorbidity profiles were identified in the post-AMI period: Acute cardio-renal-respiratory instability with chronic metabolic disease (ACUTE-CARD), Cardiometabolic disease with mixed arrhythmic-ischemic burden (CARDIOMIX), and Smoking-related cardiovascular disease with multimorbidity (SMO-CARD). CatBoost predicted profile membership with AUROC 0.77. Participants in the SMO-CARD cluster showed the highest rates of mortality, while ACUTE-CARD had the most favourable outcomes (SMART risk score = 11.2, and 6.8% CVD deaths). SMO-CARD displayed a broad range of cardiopulmonary and systemic associations. PheWAS revealed profile-specific genetic associations and pathway enrichments were consistent with clinical features; for example, cardiometabolic genes were associated with the CARDIOMIX cluster, and immune-related pathways were associated with SMO-CARD, supporting the biological plausibility of these profiles.

**Conclusion:** Integrating temporal clustering with explainable machine learning reveals distinct multimorbidity patterns in AMI patients. This framework supports personalised risk stratification and outcome prediction in clinical care.

## 1. Introduction

Acute myocardial infarction (AMI), commonly referred to as a heart attack, is a leading cause of mortality and disability worldwide^1–3^. It is a manifestation of ischaemic heart disease (IHD), which results from reduced or obstructed blood flow to the heart muscle, leading to tissue damage and thus significant morbidity and mortality. Globally, ischaemic heart disease is responsible for approximately nine million deaths annually^4^, nearly half of which are attributable to myocardial infarction (MI)^5^. Over half of post-MI patients die within five years of their initial AMI. Among those aged 65–79, life expectancy reductions of 56% are reported, with even steeper declines (62–80%) observed in patients aged 80 or older^6^. The global burden of IHD continues to rise, driven by ageing populations and the increasing prevalence of metabolic and lifestyle-related risk factors^4^. In the UK, an estimated 2.3 million people live with coronary heart disease, with 68,000 deaths attributed annually^7^. While age-standardised IHD death rates have declined in high-sociodemographic index (SDI) countries due to improved healthcare access, risk factor management, and preventative measures^8–11^, rates have increased in low- and middle-SDI countries^12^. These disparities are linked to differences in controlled metabolic risks, such as fasting plasma glucose, Body Mass Index (BMI), smoking prevalence, and exposure to environmental pollutants^13^.

AMI patient readmission rates within 30 days of treatment range from approximately 11 to 14%, often driven by cardiac factors such as acute coronary syndrome, angina, ischemic heart disease, and heart failure. It is important to clarify that “acute myocardial infarction” in this context may refer to both initial and recurrent events; however, the specific contribution of recurrent AMI to readmission rates requires further specification^14^. In the last 20 years, interventions such as reperfusion, primary percutaneous coronary intervention (PCI), dual antiplatelet therapy, statins, beta-blockers, and ACE inhibitors/ARBs have significantly improved survival and reduced recurrent ischemic events, stroke, and heart failure in patients with ST-elevation myocardial infarction (STEMI)^15^. Despite these advances, event rates have plateaued in recent years, underscoring the need for new approaches to improve long-term outcomes. This is particularly pertinent in the context of multimorbidity – the coexistence of multiple conditions and diseases – which is increasingly common among AMI patients and complicates both treatment and prognosis^16,17^. For example, common comorbidities as risk factors for 30-day readmission included kidney disease, heart failure, COPD (chronic obstructive pulmonary disease), and diabetes mellitus^18^. The number of comorbidities in post-MI patients is associated with higher mortality rates and is more often due to cardiovascular (CV) disease than non-CV^19^.

Traditional methods to analysing AMI in the context of multimorbidity focus on risk assessment through indices such as the Age-Adjusted Charlson Comorbidity Index (ACCI), or Health-Related Quality of Life (HRQoL). The study of Wei et.al^20^ mapped multimorbidity-weighted index (MWI) conditions to ICD-10 codes, thus expanding them to develop a new MWI-ICD10 and updated MWI-ICD9. This showed the MWI-ICD10 captured chronic condition prevalence nearly identical to MWI-ICD9, enabling consistent quantification of disease burden. These studies, however, often rely on simplistic metrics, such as counting the number of comorbidities, without addressing the complex interrelationships between such conditions or their temporal progression and accumulation^21^. For example, Simard et al.^22^ systematically reviewed 22 multimorbidity measures using ICD codes from health administrative data, assessing their robustness and validation processes. Their findings revealed significant variability in the measures, with about one-third rated as low to moderate quality, highlighting the need for standardisation. These methods are complemented by technologies such as machine learning (ML) and multi-omics, which offer potential enhancements in predicting AMI outcomes and improving patient care^23–26^. The integration of ML techniques in the study of AMI has led to significant advancements in identifying biomarkers and improving diagnostic accuracy^23,24,27^. Some data-driven approaches to multimorbidity have leveraged techniques such as Latent Class Analysis (LCA), an unsupervised machine learning and statistical method to identifying clusters and patterns of disease, and utilised biomarkers for prognosis^28^. In other examples, clustering techniques such as KMeans have been effective in analysing AMI disease progression, while explainable AI models such as XGBoost have enhanced the interpretability of AMI predictions^29–31^.

While these studies have contributed valuable insights into advancing AMI diagnosis and prediction, they primarily used static snapshots of patient data, focused on a limited range of AMI comorbidities, and do not consider dynamic interactions that drive disease progression. This fragmented approach hinders the identification of high-risk patient subgroups, prediction of adverse outcomes, and development of targeted prevention or treatment strategies. To overcome these limitations, our study employed advanced methodologies to analyse multimorbidity patterns and their temporal evolution in participants from the UK Biobank dataset who had suffered AMI. By integrating unsupervised Dynamic Time Warping (DTW) clustering^32^, topic modelling of cluster-specific diagnoses, supervised prediction models, explainability techniques, network interaction visualisations, Phenome-Wide Association Studies (PheWAS), and survival analysis, we aimed to extract temporal patterns, identify consistent multimorbidity associations, and uncover the complex interplay of disease trajectories in AMI. This approach addresses critical gaps in understanding and managing AMI-associated multimorbidity and can be extended to other areas of unmet clinical need.

## 2. Methods

### 2.1 Data source and study participants

Data from the UK biobank (accessed under application number: 83988) of patients’ diagnosis with acute myocardial infarction (ICD-10 code: I21), and associated health conditions were obtained. The UK Biobank is a large-scale biomedical database and research resource established in 2006. It involves approximately 500,000 participants aged 40-69 years at recruitment^33^. Participants underwent extensive assessments, including physical measurements, medical history, lifestyle questionnaires, and biological samples (blood, urine, and saliva). The extracted dataset further contains participants’ socio-demographic information (sex and ethnicity, risk factors (age, waist and hip circumference, and Body Mass Index (BMI), Smoking status), clinical outcomes (All-cause Mortality), as well as the Index of Multiple Deprivation (IMD) scores. The IMD scores are based on the UK government’s qualitative studies of deprived areas within the local councils of England, Wales and Scotland, published during the UK Biobank baseline visits, which took place between 2006 and 2010^34^. The final dataset includes 12,701 participants, diagnosed with AMI, after recruitment to UK Biobank.

#### 2.1.1 Data pre-processing

Each of 12,701 participants included in the analyses had additional diagnoses captured by ICD-10 codes. We refined the ICD-10 codes to focus on long-term or chronic conditions and comorbidities, resulting in 1,008 conditions after excluding cases related to pregnancy and childbirth (O04-O99, Z32-Z61), perinatal conditions, accidents (V09-V97), unclassified categories, general symptoms such as COVID-19 (P28), other non-specific symptoms (R09-R68), and personal histories of certain conditions (Z00–Z98). Very rare ICD-10 codes with fewer than five occurrences in the cohort were also excluded. The “icd10-cm” package (version 0.0.5) was used to map ICD-10 codes to the relevant condition names. We divided the dataset into two periods: the pre-AMI period (diagnoses recorded between 365 days to 0 days prior to the date of AMI diagnosis) and the post-AMI period (diagnoses recorded within 0–5 years following AMI diagnosis). All participants had at least one pre-AMI and post-AMI diagnosis. For both pre- and post-AMI diagnoses, we calculated their total accumulated ICD-10 (diagnoses) count, the disease progression rate, and the time to the first diagnosis during this period.

All analyses were performed using Python (version 3.8.10). The analyses and exploration were carried out in Jupyter Notebook (version 5.2.0).

### 2.2 Dynamic time warping clustering of AMI participants’ ICD-10 codes data

To investigate disease-trajectory patterns during the post-AMI period, we constructed, for each participant, a temporal sequence of diagnoses based on the recorded age at which each diagnosis occurred. In the dataset, each ICD-10 diagnosis was represented as a column populated with the participant’s age at diagnosis (or left blank if absent). Temporal ordering was therefore determined by sorting diagnoses according to these recorded ages, allowing us to capture both the order and cumulative count of post-AMI diagnoses without relying on precise date stamps.

The resulting sequences were padded (i.e., shorter diagnosis sequences were extended with placeholder values such as zeros) to a uniform length based on the longest sequence in the dataset and standardised using the *TimeSeriesScalerMeanVariance* method to ensure consistency across samples. The “tslearn” package (version 0.6.3) was used for TimeSeriesKmeans Dynamic Time Warping analysis to identify patterns in early diagnosis trajectories, we applied *TimeSeriesKMeans* clustering with DTW as the distance metric. In this context, *k* refers to the number of participant clusters (i.e., grouping patients based on similarities in their temporal diagnosis patterns), analogous to the standard k-means algorithm but adapted to use DTW instead of Euclidean distance. We evaluated clustering performance across a range of values (k = 2 to 9) using the Elbow method (based on within-cluster sum of squares), Silhouette score, and Davies–Bouldin index to determine the optimal number of clusters. All clustering was conducted with a fixed random seed (42) to ensure reproducibility. The resulting cluster labels were applied to the same participants in the pre-AMI dataset, enabling post-clustering analyses to identify clinically meaningful patterns and predict each participant’s cluster trajectory.

### 2.3 Topic Modelling of Cluster-Specific Diagnoses

To characterise the diagnostic themes within each cluster, we applied Latent Dirichlet Allocation (LDA) to the binary presence of ICD-10 codes. For each cluster, ICD-10 codes were treated as “words” within a document, and LDA was used to extract two dominant topics per cluster. To support clinical interpretation of these topics, we examined the top fifty ICD-10 codes with the highest topic probabilities and mapped them to broader clinical categories using a predefined dictionary. This dictionary was constructed by: (i) using ICD-10 chapter and block headings as the primary organisational structure; (ii) refining category boundaries to ensure clinical coherence (e.g., grouping related cardiometabolic diagnoses such as hypertension, hyperlipidaemia and type 2 diabetes); and (iii) validating code assignments through review by domain experts. Based on the LDA-derived themes and clinical category distributions, clusters were labelled as follows:

1. ACUTE-CARD: Acute cardio-renal-respiratory instability with chronic metabolic disease.
2. CARDIOMIX: Cardiometabolic disease with mixed arrhythmic-ischemic burden
3. SMO-CARD: Smoking-related cardiovascular disease with multimorbidity

A heatmap was generated to visualise the frequency of clinical categories across cluster, highlighting dominant disease patterns and aiding in the interpretation of cluster phenotypes (see Figure 2).

**Figure 1:**
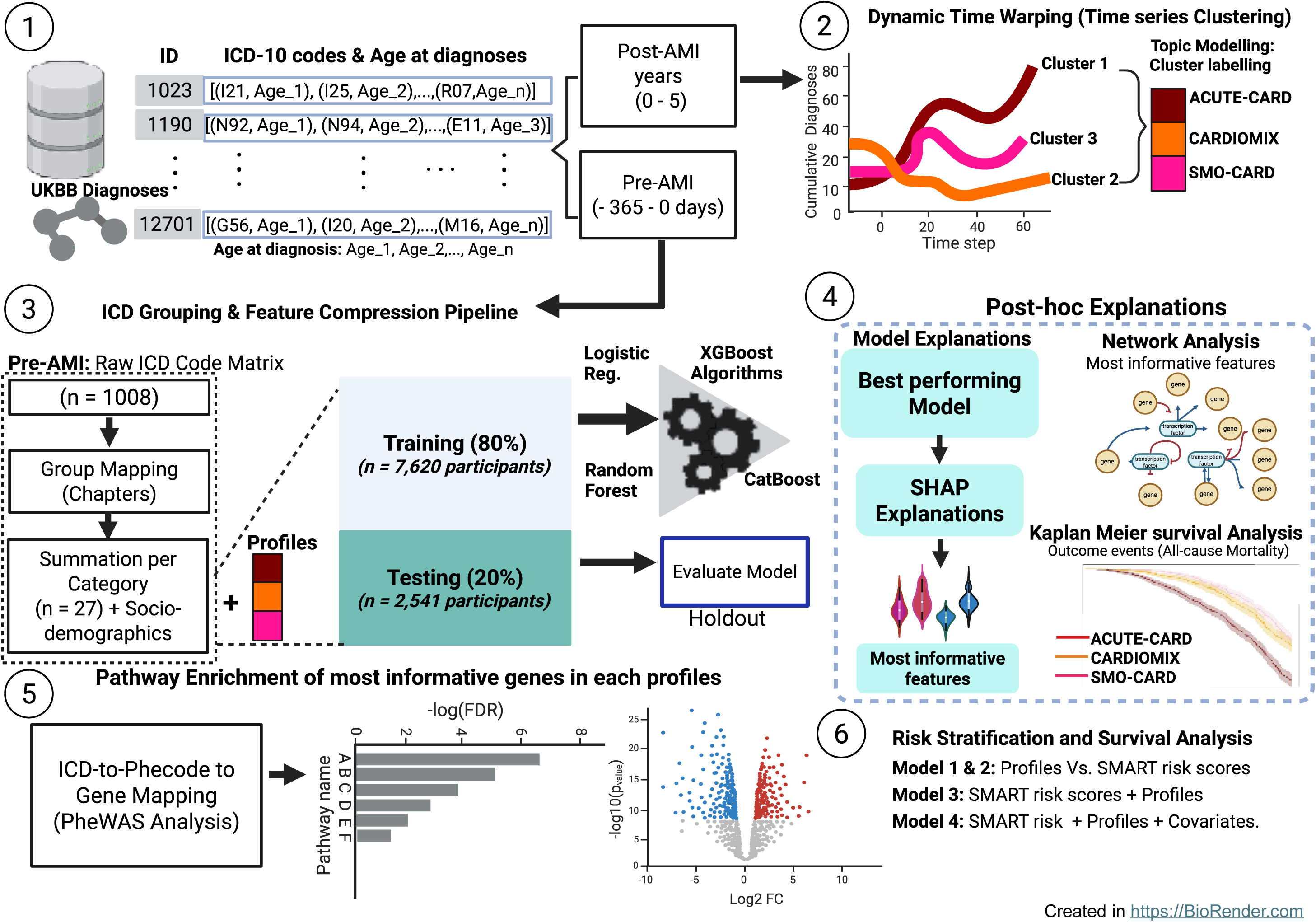
Schematic representation of Study workflow and Computational Pipeline. (1) Data collection: UK Biobank data were collected for 12,701 participants with an AMI diagnosis, inclusive of 1,008 different ICD-10 codes. The dataset is structured with participants as rows, ICD-10 codes as columns, and the ages at diagnosis recorded in the cells. Absence of a diagnosis is represented as NaN. Using these data, we created a sequence of diseases for each participant to facilitate downstream analyses. We study the data from two angles: pre-AMI (diagnoses received at no more than 365 days prior to AMI diagnosis) and Post-AMI Period (diagnoses received in the 0–5 years following AMI diagnosis). In the post-AMI dataset, we performed (2) Time Series Analysis: each participant’s disease sequence was constructed by ordering diagnoses according to the recorded age at which they occurred. These age-ordered sequences were then padded to a uniform length and used as input for dynamic time warping K-Means clustering. We further perform a thematic topic modelling of ICD codes in each cluster to derived meaningful labels (profile) for each cluster which were mapped back to the ICD dataset of pre-AMI diagnoses for further analysis. (3) Feature matrix was created using the pre-AMI dataset with profile membership as target labels into 80% training and 20% testing datasets. (4) Supervised learning models: Four supervised learning algorithms - Logistic Regression, Random Forest, CatBoost, and XGBoost - were trained on the pre-AMI dataset. CatBoost outperformed the other models and was selected for further evaluation. Post-Hoc Model Interpretation and Analysis: Post-hoc SHapley Additive exPlanations (SHAP) analysis was conducted to interpret the model’s predictions. The most important features identified by SHAP were visualised through network analysis. Additionally, survival analysis was performed to evaluate the clinical outcome – composite event (Myocardial Infarction, Stroke, CVD deaths, and All-cause Mortality), for each multimorbidity profile in the testing set. (5) ICD codes identified from non-zero SHAP values were mapped to Phecodes from the PheWAS study, and pathway enrichment analysis was conducted to identify biological processes associated with the health conditions for each profile. (6) Survival analysis was conducted to assess the risk stratification ability of the profile, SMART risk scores, and conventional covariates.

**Figure 2:**
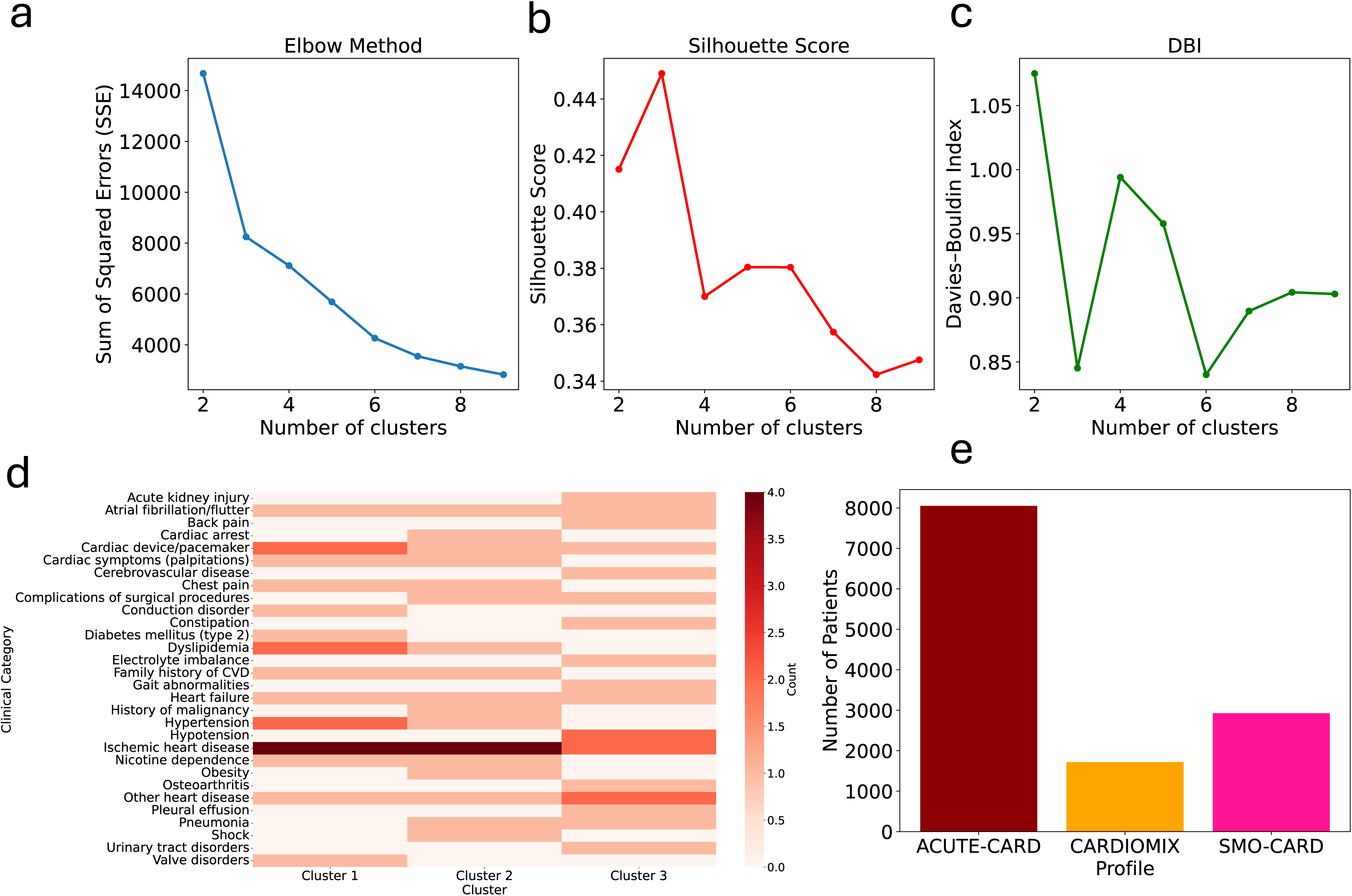
AMI participants profiles from clustering their ICD-10 codes and age at diagnoses Panel A: Elbow Method used to identify the optimal number of clusters by observing the SSE “elbow” point. Panel B: Silhouette Score for evaluating the DTW cluster cohesion and separation. Panel C: DBI used for measuring intra-cluster similarity and inter-cluster difference (lower values are better). Panel D illustrates a heatmap from thematic topic modelling of cluster-specific diagnoses. Panel E presents a bar chart showing the number of AMI participants in each profile. Profiles – ACUTE-CARD (Cluster 1) was colour-coded dark red, CARDIOMIX (Cluster 2) was colour-coded orange, and SMO-CARD was colour-coded pink (Cluster 3).

### 2.4 Statistical analysis

Following the identification of distinct clusters (profiles) using the post AMI diagnoses, we compared baseline characteristics across the three groups. To evaluate the distribution of data, the Kolmogorov–Smirnov test was applied. Given that continuous variables did not adhere to a normal distribution, we utilised the nonparametric Kruskal–Wallis test for their comparison. Differences in categorical variables were assessed using the χ² test, with Bonferroni correction applied for multiple comparisons where appropriate. Chi-squared tests with corrections were performed to assess statistical associations in the prevalence of ICD-10 code-defined diseases across profiles. A threshold of *p-value < 0.05* was used as the criterion for significance. To mitigate false positives arising from multiple comparisons, we applied the Benjamini–Hochberg correction for categorical variables and the Bonferroni correction for continuous variables. Descriptive statistics were reported as median (25^th^, 75^th^ percentile) for continuous variables and as count (percentages) for categorical data (See Table 1). The “scipy” package (version 1.7.3) was employed for descriptive statistics and statistical tests.

**Table 1:** Baseline characteristics of AMI participants stratified by multimorbidity profile.

| Attributes | Whole AMI cohort<br>(n = 12701) | ACUTE-CARD<br>(n = 8052) | CARDIOMIX<br>(n = 1720) | SMO-CARD<br>(n = 2929) | p-value |
| --- | --- | --- | --- | --- | --- |
| Sex (Female) (%) | 8778 (69.1) | 5651 (70.2) | 1202 (69.9) | 1925 (65.7) | <0.0001 |
| Age (Years) | 62.0 (56.0, 66.0) | 61.0 (55.0, 65.0) | 61.0 (55.0, 65.0) | 64.0 (60.0, 67.0) | <0.0001 |
| BMI | 28.0 (25.4, 31.1) | 27.8 (25.3, 30.7) | 27.7 (25.2, 30.8) | 28.6 (25.8, 32.2) | <0.0001 |
| IMD score | 13.8 (7.5, 26.0) | 13.5 (7.2, 24.8) | 12.7 (6.9, 24.3) | 15.6 (8.4, 29.8) | <0.0001 |
| Waist Circumference | 96.0 (88.0, 104.0) | 95.0 (87.0, 103.0) | 95.0 (87.0, 103.0) | 98.0 (89.0, 107.0) | <0.0001 |
| Hip Circumference | 103.0 (98.0, 109.0) | 103.0 (98.0, 108.0) | 103.0 (98.0, 109.0) | 104.0 (99.0, 111.0) | <0.0001 |
| Stroke (%) | 821 (6.5) | 357 (4.4) | 90 (5.2) | 374 (12.8) | <0.0001 |
| CVD Death (%) | 1224 (9.6) | 546 (6.8) | 203 (11.8) | 475 (16.2) | <0.0001 |
| All-cause Mortality (%) | 2673 (21.0) | 1034 (12.8) | 354 (20.6) | 1285 (43.9) | <0.0001 |
| Composite CVD (%) | 435 (3.4) | 200 (2.5) | 70 (4.1) | 165 (5.6) | <0.0001 |
| SMART Risk Score | 11.8 (9.1, 15.2) | 11.2 (8.9, 14.2) | 11.2 (8.8, 14.8) | 14.0 (10.4, 18.4) | <0.0001 |
| SMART REACH Score<br>CVD Mortality | 2.5 (2.0, 3.2) | 2.4 (2.0, 3.1) | 2.3 (1.8, 2.9) | 2.8 (2.2, 3.8) | <0.0001 |
| SMART REACH Score<br>All-cause Mortality | 0.6 (0.2, 0.9) | 0.5 (0.2, 0.8) | 0.5 (0.2, 0.8) | 0.8 (0.4, 1.2) | <0.0001 |
| SBP (mmHg) | 144.0 (132.0, 156.0) | 144.0 (132.0, 156.0) | 142.0 (130.0, 156.0) | 144.0 (132.0, 157.0) | 0.027 |
| DBP (mmHg) | 84.0 (77.0, 91.0) | 85.0 (78.0, 92.0) | 84.0 (77.0, 91.0) | 82.0 (74.0, 90.0) | <0.0001 |
| Pulse Rate (BPM) | 69.0 (62.0, 78.0) | 69.0 (62.0, 77.0) | 68.0 (61.0, 77.0) | 69.0 (61.0, 78.0) | 0.182 |
| eGFR (mL/min/1.73 m2) | 90.0 (80.0, 100.0) | 91.0 (81.0, 101.0) | 91.0 (81.0, 100.0) | 85.0 (73.0, 96.0) | <0.0001 |
| Diabetes (%) | 853 (6.7) | 280 (3.5) | 113 (6.6) | 460 (15.7) | <0.0001 |
| CAD (%) | 1235 (9.7) | 318 (3.9) | 254 (14.8) | 663 (22.6) | <0.0001 |
| CD (%) | 195 (1.5) | 78 (1.0) | 23 (1.3) | 94 (3.2) | <0.0001 |
| AAA (%) | 29 (0.2) | 9 (0.1) | 1 (0.1) | 19 (0.6) | <0.0001 |
| PAD (%) | 179 (1.4) | 46 (0.6) | 19 (1.1) | 114 (3.9) | <0.0001 |

### 2.5 Supervised Multi-classification

A feature matrix using ICD-10 codes from the pre-AMI data socio-demographic variables (age, IMD score, BMI, and sex) was constructed, with the clustering profiles used as the target labels. A structured feature engineering process, aimed at improving interpretability and reducing dimensionality issues, was applied. ICD codes were transformed into a binary present/absent ‘1’ or ‘0’. Next, individual ICD codes were grouped into the 27 organ systems and ICD-10 Chapters^35^. This grouping reduced the feature space from over 1000 codes to manageable set of interpretable variables. For each participant, we calculated the total number of diagnoses within each category, resulting in a count-based feature matrix. The feature matrix was then scaled by standardisation.

The feature matrix was split into training (80%) and testing (20%) sets. To build a multiclass classifier to predict profile membership four supervised learning algorithms were employed - Logistic Regression, Random Forest, XGBoost, and CatBoost. Hyperparameter optimisation was performed using GridSearchCV with 3-fold cross-validation, optimising for weighted F1-score. To address class imbalance, custom class weights were applied, with increased penalties for misclassifying the CARDIOMIX group, as it represented the smallest class and is associated with higher clinical complexity and adverse cardiovascular risk.

Model performance was evaluated on the testing (20%) set. The area under the curve (AUC) of the receiver operating characteristic (ROC) curve (i.e., Multiclass ROC-AUC (one-versus rest)) was calculated (See Supplementary Table 2). In addition, indicators weighted average accuracy, precision, recall, and confusion matrices were utilised to analyse the performance of different classifiers^36,37^. SHAP (SHapley Additive exPlanations) was used to explain the output of machine learning models^38^ by illustrating how and to what extent each feature contributes to the result^31^. SHAP values were computed for each feature across all test samples, enabling identification of the topmost influential features per profile membership. Supervised learning tasks were conducted using the “scikit-learn” package (version 1.3.2).

### 2.6 Network analysis of disease co-occurrence

To gain deeper insight into how specific health conditions co-occur within individuals across the predicted profiles, we constructed a feature correlation network using Pearson correlation coefficients among fifty top ICD codes from LDA analysis. Using the post-AMI cohort, we generated a co-occurrence matrix quantifying how often each pair of feature categories appeared together in individual records. This matrix was then used to build an undirected, weighted graph in which nodes represented feature categories and edges indicated co-occurrence, weighted by their frequency across participants. Only edges with an absolute correlation above the 95th percentile were retained. Node attributes included correlation coefficient (weights) and degree centrality. To visualise the disease interaction landscape, the network was imported into Cytoscape (v3.10) for complex network analysis.

### 2.7 Risk Stratification and Survival Analysis of Adverse Clinical Outcomes

Risk across the profiles was calculated using stablished secondary prevention tools using the SMART (Second Manifestations of Arterial Disease) risk score^39^. The resulting SMART risk model is used to provide a more comprehensive and accurate estimation of the 10-year risk and lifetime risk for recurrent Major Adverse Cardiovascular Events (MACE) in patients with existing Atherosclerotic Cardiovascular Disease (ASCVD). Required variables systolic and diastolic blood pressure, pulse rate, estimated glomerular filtration rate (eGFR), diabetes, coronary artery disease, cerebrovascular disease, peripheral artery disease, and abdominal aortic aneurysm were obtained from UK Biobank AMI baseline data, alongside years since first cardiovascular diagnosis. Scores were expressed as risk percentages.

An outcome of all-cause mortality event was considered for this analysis. Time-to-event variables were calculated from the UK Biobank baseline to the event date within five years, with follow-up censored at the earliest event. Cox proportional hazards models were fitted sequentially: Model 1: (profiles only), Model 2 (SMART risk score), Model 3 (profiles + SMART risk score), and Model 4 (profiles + SMART risk score + conventional covariates: age, sex, IMD score, systolic blood pressure, pulse rate, eGFR, diabetes, coronary artery disease, cerebrovascular disease, and peripheral artery disease). Hazard ratios (HRs) and 95% confidence intervals (CIs) were reported. Models’ discrimination was evaluated using Harrell’s concordance index (C-index) and competing-risk Kaplan–Meier curves were generated across profiles. Analyses were conducted in Python (v3.10) using the *lifelines* package.

### 2.8 PheWAS analysis and pathway enrichment

To explore potential biological mechanisms and pathways associated with the identified profiles, a Phenome wide association study (PheWAS) was conducted. The most influential health conditions identified during the thematic topic modelling of profile-specific diagnoses were mapped to phenotype codes (phecodes) using the PhecodeX (Extended) version 1.0 resource from the PheWAS catalogue^40–42^.

For each mapped phecode, PheWAS analysis was carried out using a dataset of SNP-phenotype associations derived from the study cohort. The dataset was filtered by Phecode to identify SNPs with significant associations (p-value < 0.05). SNPs were sorted by p-value to prioritise the most statistically significant associations between genes and each profile; associated gene names, odds ratios, and GWAS annotations were further extracted (see Supplementary tables 3 – 6). Significant SNPs were next used for pathway enrichment analysis using the Reactome Pathway Database^43^. Associations for the significant SNPs for each profile were further visualised (see Supplementary Figure 2 – 4).

## 3. Results

### Demographic characteristics of the study participants

As detailed in the Methods section, data from UK Biobank participants (502,000 in total) were analysed for the occurrence of an AMI and 12,701 participants were found to have had such an event recorded. The characteristics of these participants is recorded in Table 1 inclusive of some specific clinical features recorded on enrolment into the cohort. The median (25th to 75th percentile) age at baseline for the overall cohort was 62 (56, 66) years, while the median age at AMI diagnosis was 69 (63, 74) years. The cohort comprised 30.9% males and 69.1% females, 88.9% are White British, reflective of the UK Biobank cohort. The profiles exhibited distinct characteristics across socio-demographic and outcome variables (see Table 1). Participants had a median of 8.0 ICD-10 diagnoses (Range 0.0 - 66.0) in the post-AMI period, and a median of 4.0 (range 0.0 - 47.0) in the pre-AMI period.

### 3.1 Distinct Temporal Patient Profiles

To determine the optimal number of DTW cluster partitions, we evaluated clustering performance using the Elbow Method, Silhouette Score, and Davies-Bouldin Index. As shown in Figure 2 (A through C). The results indicated that the optimal number of clusters is three, providing the most appropriate balance across all metrics with a Silhouette Score of 0.45 and a Davies-Bouldin Index of 0.85 (lower the better), indicating a well-separated structure within the clusters.

To explore the dominant diagnostic themes within each cluster, we applied LDA analysis to identify co-occurring diagnoses that define the characteristics of each group (see Supplementary Table 2). The three resulting profiles are as follows: a) ACUTE-CARD with 8,052 participants. b) CARDIOMIX with 1,720 participants. C) SMO-CARD with 2,929 participants (Figure 2). Profile labels were mapped to participants’ pre-AMI diagnosis data for supervised prediction of profile membership and all subsequent analyses.

#### 3.1.1. Profile 1: Acute Cardiorenal–Respiratory with Chronic Metabolic Disease (ACUTE-CARD)

This profile is characterised by a combination of chronic cardiometabolic disease and acute physiological instability. The dominant diagnostic themes identified by LDA included dyslipidaemia, cardiac conduction abnormalities, atrial fibrillation or flutter, hypertension, heart failure, non-specific cardiac disease, and chronic ischaemic heart disease. These features suggest a pattern of recurrent cardiovascular strain, likely arising from a long-standing burden of vascular and metabolic risk. Additional themes in this group included type 2 diabetes, nicotine dependence, and chest pain presentations. The frequent appearance of chronic ischaemic codes (I20, I24 and I25) highlights a group with substantial underlying coronary disease susceptibility. Recurrent arrhythmia, conduction disturbance and heart failure codes point to electrical and structural cardiac instability as a key characteristic of this profile.

Demographically, participants were mostly female (70.2 percent) with a median age of 61 years (interquartile range 55 to 65 years). Socioeconomic deprivation was moderate (median IMD score 13.5). The group had a median BMI of 27.8 kg/m² and waist circumference of 95 cm. Cardiovascular comorbidities included diabetes (3.5 percent), coronary artery disease (3.9 percent), and peripheral artery disease (0.6 percent). Despite the considerable burden of chronic cardiovascular pathology, mortality was comparatively low with all-cause mortality at 12.8 percent, although the cluster demonstrated elevated vascular risk as reflected in the SMART and SMART-REACH scores. The combination of electrical, metabolic and ischaemic instability indicates a population with cumulative cardiac stress and reduced physiological reserve.

#### 3.1.2. Profile 2: Cardiometabolic with Mixed Arrhythmic–Ischaemic Burden (CARDIOMIX)

The CARDIOMIX profile comprises individuals with both metabolic risk factors and a combination of arrhythmic, ischaemic and respiratory instability. The LDA themes for this cluster included cardiac arrest, arrhythmias, ischaemic heart disease, acute kidney injury, dyslipidaemia and prior vascular risk represented by family history of cardiovascular disease. Additional themes included pneumonia, hypotension, obesity, nicotine dependence and atrial fibrillation. This mixture reflects a pattern of cardiometabolic disease where metabolic dysfunction coexists with arrhythmic risk and episodes of acute physiological compromise. The presence of sepsis-related or respiratory codes in particular themes may indicate susceptibility to decompensation during intercurrent illness. Hypertension and ischaemic heart disease appeared consistently, reinforcing the presence of chronic vascular strain. Participants were mostly female (69.9 percent) with a median age of 61 years (interquartile range 55 to 65 years). Members of this profile had a BMI of 27.7 kg/m², waist circumference of 95 cm and were the least socioeconomically deprived of the three clusters (median IMD 12.7). Diabetes was present in 6.6 percent and coronary artery disease in 14.8 percent. All-cause mortality was moderate at 20.6 percent and composite cardiovascular outcomes occurred in 4.1 percent. Although the CARDIOMIX profile showed fewer extreme metabolic or respiratory disturbances than ACUTE-CARD, SMART and SMART-REACH risk scores indicate a sustained level of vascular risk that remains clinically significant.

#### 3.1.3. Profile 3: Smoking-Related Cardiovascular Disease with Multimorbidity (SMO-CARD)

The SMO-CARD profile is distinguished by multimorbidity strongly linked with smoking and its systemic effects. LDA themes for this cluster included conduction disorders, non-specific cardiac disease, chronic cerebrovascular syndromes, urinary tract disorders, musculoskeletal pain, mobility difficulties and chronic constipation. Additional themes captured as part of the second topic included nicotine dependence, atrial fibrillation, pleural effusion, electrolyte imbalance, pneumonia, acute kidney injury and heart failure. This broad array of respiratory, renal, musculoskeletal and cardiovascular codes illustrates a diffuse multimorbidity pattern consistent with long-term smoking exposure. Electrical cardiac instability and recurrent respiratory conditions were frequent, supporting the interpretation of a group with systemic frailty and heightened vulnerability to acute illness. The presence of pleural effusion and pneumonia further underlines the respiratory contribution to their disease trajectory. Participants had a higher median age of 64 years (interquartile range 60 to 67 years) and demonstrated greater socioeconomic deprivation (median IMD 15.6). BMI was 28.6 kg/m² and waist circumference was 98 cm, the highest of the three clusters. Cardiovascular comorbidities were more prevalent than in the other groups, including diabetes (15.7 percent), coronary artery disease (22.6 percent), cerebrovascular disease (3.2 percent) and peripheral artery disease (3.9 percent). All-cause mortality was high at 43.9 percent and cardiovascular mortality was also the most pronounced in this cluster. The SMART-risk score was markedly elevated, reflecting the heavy cumulative burden of vascular, respiratory and metabolic disease.

### 3.2. An explainable prediction model of AMI profiles

Based on 27 feature-categories of ICD-10 codes assigned to participants in the pre-AMI period we aimed to predict profile membership by training four models: Logistic Regression, Random Forest, XGBoost and CatBoost. Hyperparameters were optimised via GridSearchCV, and performance was evaluated on a 20% hold-out testing set. Weighted average metrics are summarised in Supplementary Table 2.

CatBoost achieved the highest performance among the test models tested, with a test accuracy of 65%, F1-score of 0.61, and AUROC of 0.77, indicating moderate effectiveness in capturing complex patterns in predicting profile membership of AMI participants based on pre-AMI diagnoses. Figure 3 shows the AUROC curves and the class-level confusion matrix. To better understand the prediction results of the best-performing model at a granular level, we employed the SHAP interpretation approach. In the ACUTE-CARD profile, the top five most influential ICD chapters (based on the highest SHAP values) are ‘Ischemic heart disease’, ‘Dyslipidemia & obesity’, ‘Hypertension and hypertensive heart disease (HHD)’, ‘major psychiatric disorders’, and ‘arrhythmias & conduction’ (Figure 4a). In the CARDIOMIX profile, the top five most important contributors are ‘Valvular & Myocardial disease’, ‘Atherosclerosis & Vascular’, ‘Symptoms, family history, and health status’, ‘Ischemic heart disease’, and ‘IMD score’ (Figure 5a). These features are particularly relevant in distinguishing this profile from the others. In the SMO-CARD profile, the five most significant features are ‘Age’, ‘ischemic heart disease’, ‘BMI’, ‘Other renal & genitourinary disease’, and ‘IMD score’ (Figure 4, panel (A – C)).

**Figure 3:**
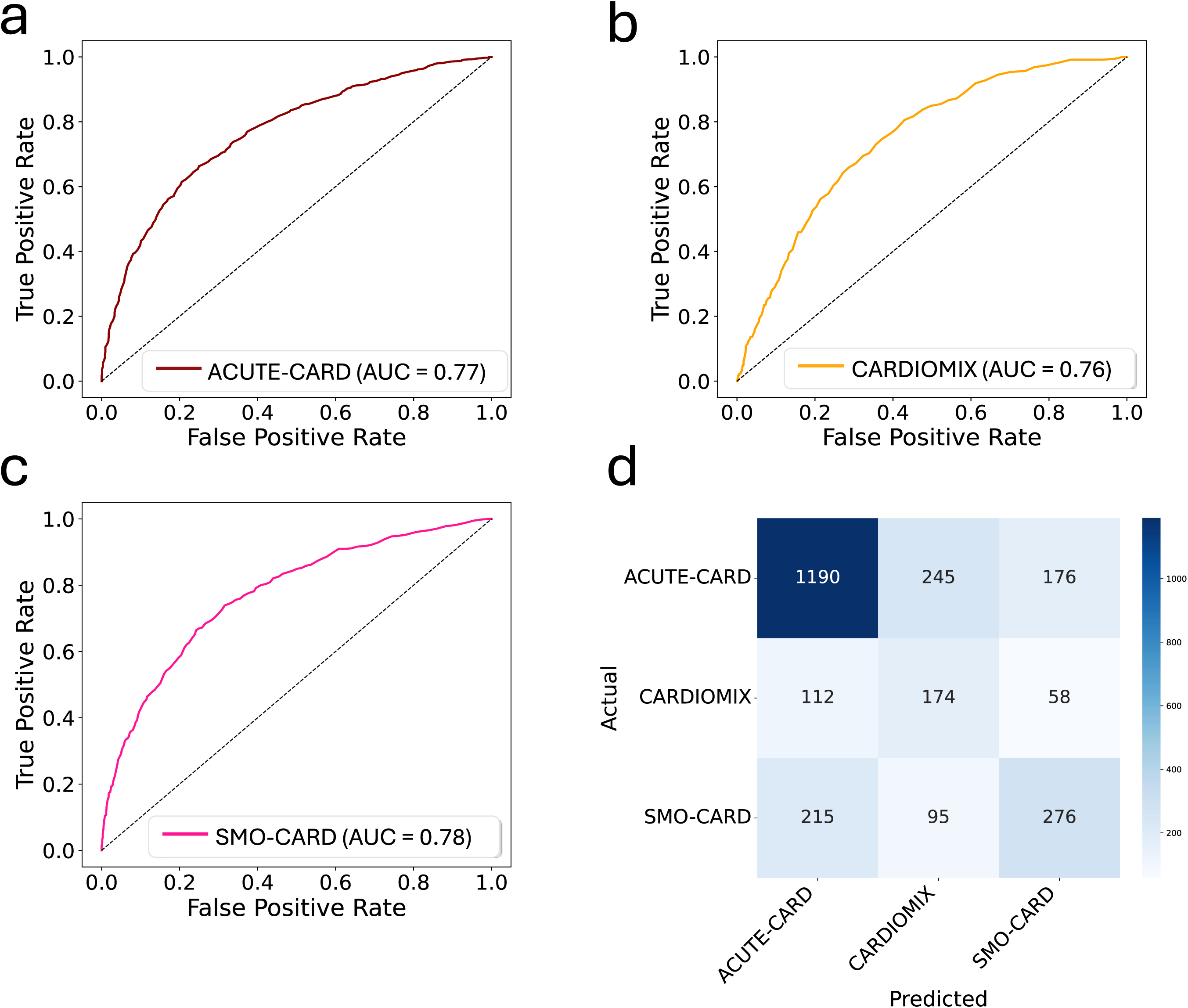
Performances of the supervised classifier for prediction of distinct ICD-10 profiles in AMI participants. Panel (a) - (c) depicts the AUROC scores for the CatBoost algorithm during testing across the profiles. AUROC scores displayed for the profiles – ACUTE-CARD (darkred), CARDIOMIX (orange), and SMO-CARD (pink).Panel (d) depicts the confusion matrix for the multiclass classification of profiles on test set using the CatBoost algorithm. The confusion matrix provides a detailed breakdown of the classification outcomes by comparing the predicted and actual labels. Each row of the matrix represents the true class, while each column corresponds to the predicted class.

**Figure 4:**
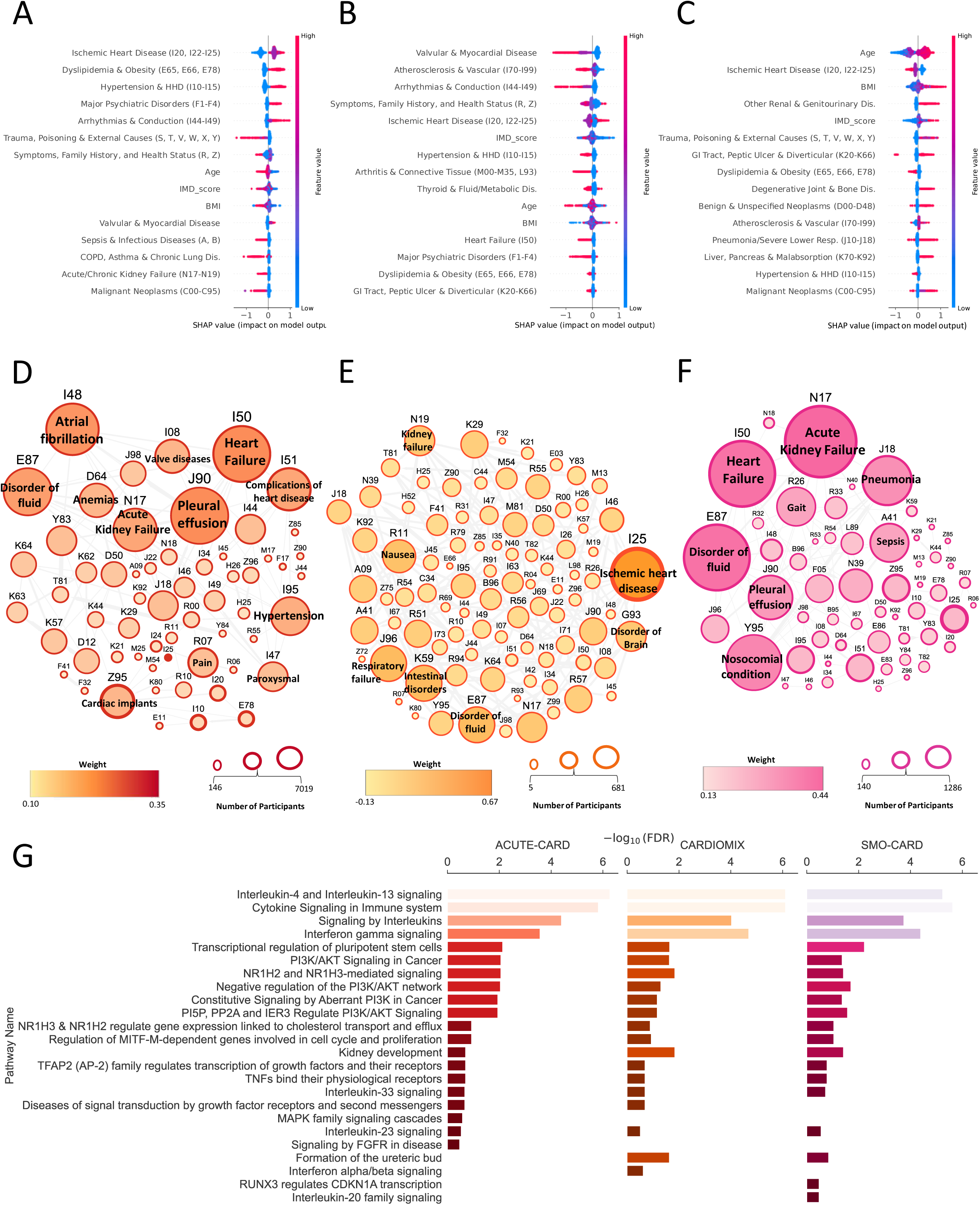
SHAP-based feature interpretation, network connectivity, and pathway enrichment across the profiles Colour bars indicate relative feature values (high to low); red points denote higher feature values and blue points denote lower values, with horizontal position reflecting SHAP impact. (A–C) SHAP summary plots for ACUTE-CARD, CARDIOMIX, and SMO-CARD showing the top 20 features ranked by mean absolute SHAP value. (D–F) Network analyses of the top 50 ICD codes associated with the ACUTE-CARD, CARDIOMIX, and SMO-CARD LDA topics. Node size reflects prevalence; edges represent co-occurrence; border thickness indicates correlated weight. (G) Pathway enrichment of the top 20 biological processes associated with the top 50 ICD codes from the dominant LDA topic, stratified by profile.

### 3.3. Disease Co-occurrence Networks

To investigate the interrelationships among the most influential diagnostic features within each profile, correlation-based feature networks were constructed using a correlation-ranked ICD-10 codes. Each node represents an ICD-code with the matched clinical name enclosed in brackets. Edges denote strong positive or negative correlations, where the absolute magnitude of the correlation coefficient exceeds the 95th percentile threshold of the correlation distribution. These networks highlight distinct multimorbidity structures underpinning each cluster (see Figures 4, panel (D – F)).

#### 3.3.2. Multimorbidity profiles, SMART-REACH and adverse outcomes

We evaluated time to all-cause mortality with 5-years using sequential Cox proportional hazards models (Supplementary Table 5). In the Cox proportional hazards model including only the multimorbidity profile score (Model 1), which quantified the underlying severity axis, each unit increment in the score was significantly associated with a 40% increased risk of death (HR = 1.40 per unit score, 95% CI 1.23–1.59, p < 0.001). Model discrimination for the profile-only model was modest (C-index = 0.59). In the SMART-risk score–only model (Model 2), as expected, higher SMART-risk scores were strongly associated with increased event risk (HR = 1.05 per unit, 95% CI 1.04–1.07, p < 0.001), with moderate discrimination (C-index = 0.65). When both predictors were included jointly (Model 3), the SMART-risk scores remained a strong independent predictor (HR = 1.05, 95% CI 1.04–1.06, p < 0.001), while the profile score also remained statistically significant (HR = 1.25, 95% CI 1.10–1.43, p = 0.001) and discrimination improved (C-index = 0.67).

In the fully adjusted model (Model 4), which included the multimorbidity profiles, SMART-risk scores, and traditional clinical covariates, the profile score was no longer statistically significant (HR = 1.12, 95% CI 0.98–1.29, p = 0.105). The SMART-risk score continued to demonstrate strong independent prognostic value (HR = 1.03, 95% CI 1.02–1.05, p < 0.001). Among additional covariates, older age, male sex, higher socioeconomic deprivation (IMD score), and diabetes were significant predictors, whereas BMI, coronary artery disease, cerebrovascular disease, and peripheral arterial disease were not independently associated with event risk. Model discrimination improved further in the fully adjusted specification (C-index = 0.69). Kaplan–Meier survival curves showed separation by predicted profile scores in the unadjusted analysis, with attenuation of differences after adjustment in line with the findings of Model 4 (see supplementary Figure 1).

#### 3.3.3. PheWAS and Pathway Enrichment Reveal Distinct Genetic and Biological Profiles

A phenome-wide association analysis was conducted across the three profiles to identify shared and profile-specific genetic susceptibility loci linked to a wide range of clinical traits. This revealed 2,839, 2,913, and 2,808 significant SNP associations for the ACUTE-CARD, CARDIOMIX, and SMO-CARD profiles, respectively (Supplementary Figure 2). While all three profiles shared strong associations with ischaemic heart disease, primarily driven by the established CDKN2B-AS1 locus (rs4977574), the remaining signals revealed marked divergence in underlying disease aetiologies.

The CARDIOMIX profile was distinguished by a pronounced cardiometabolic signature, with its strongest association observed for Type 2 diabetes, led by the TCF7L2 locus (rs7903146), alongside significant associations with atrial fibrillation and essential hypertension. In contrast, the ACUTE-CARD profile exhibited prominent non-cardiac associations extending into renal and respiratory phenotypes, including pleural effusion and hypotension, suggesting a disease process characterised by systemic inflammation and vascular instability preceding the acute event. The SMO-CARD profile, while sharing core ischaemic heart disease associations, showed enrichment for chronic inflammatory and degenerative conditions such as diverticulosis and anaemias, consistent with cumulative tissue damage associated with long-term smoking exposure.

Analysis of cluster-specific variants revealed substantial differences in genetic architecture. CARDIOMIX harboured the largest number of unique associations, compared with markedly fewer in ACUTE-CARD and SMO-CARD (See Supplementary Figure 3). The phenotypic associations driven by these unique variants further reinforced distinct aetiologies, implicating lipid and hepatic pathways in ACUTE-CARD, neurological or complement-related processes in CARDIOMIX, and frailty- and obesity-related traits in SMO-CARD.

The pathway enrichment analysis highlights marked biological divergence between the three profiles (Figure 4g), with shared immune activation masking fundamentally distinct regulatory programmes. All profiles exhibit strong enrichment for immune signalling pathways, particularly Cytokine Signalling in the Immune System and Interferon gamma signalling, indicating that immune activation is a common backdrop to acute myocardial infarction. Against this shared baseline, the CARDIOMIX profile emerges as the most immunometabolically dysregulated, showing the strongest overall enrichment, driven by prominent Interleukin-4 and Interleukin-13 signalling alongside PI3K/AKT pathway activation and its negative regulators. The ACUTE-CARD profile shares elements of this immune–stress response but displays comparatively attenuated enrichment, consistent with a more acute, systemic inflammatory phenotype. In contrast, the SMO-CARD profile is clearly distinct, lacking dominant PI3K/AKT signatures and instead showing selective enrichment for RUNX3-regulated CDKN1A transcription and interleukin-20 family signalling, pointing towards alternative regulatory mechanisms linked to chronic tissue remodelling and inflammatory adaptation.

## 4. Discussion

This study presents a novel, data-driven framework for dissecting the intricate temporal evolution of multimorbidity following acute myocardial infarction. By integrating Dynamic Time Warping with a large-scale, longitudinal cohort, we move beyond static assessments to identify clinically distinct patient profiles defined by their dynamic disease trajectories and then molecular characteristics. The emergence of three robust multimorbidity profiles: ACUTE-CARD, CARDIOMIX, and SMO-CARD validates our methodology and provides a powerful new lens for stratifying AMI patients. This approach directly addresses a critical gap in medical research, where the sequential and interactive nature of comorbidities is often oversimplified ^21,22,44^.

Our findings reveal a profound divergence in clinical outcomes across these profiles, with the SMO-CARD profile exhibiting an increased vulnerability. This group, characterised by advanced systemic disease and functional decline, had a slightly higher risk of CVD deaths. A critical finding concerns the prognostic contribution of our clustering-derived multimorbidity profiles relative to established risk stratification tools. While the multimorbidity profile was independently associated with mortality in the unadjusted model and remained statistically significant alongside the SMART-risk^39^ score in Model 3, this predictive signal was not maintained in the fully adjusted specification Model 4. While overall Model 4 performed better, the loss of significance suggests that the prognostic information, as it relates to mortality specifically in this cohort, captured by the novel profiles is largely redundant with or mediated through the traditional clinical covariates included in the final model (namely, older age, male sex, IMD score, and diabetes).

This underscores the continued primacy of established cardiovascular risk scores and readily available clinical factors, such as SMART-risk scores, in predicting long-term mortality risk. Nevertheless, the strength of this work lies in the profiles’ ability to serve as a robust framework for aetiological stratification and phenotyping the multisystemic fragility that characterises participants post-AMI.

Furthermore, the integration of explainable machine learning and network analysis provides a transparent and interpretable basis for these clinical distinctions^45–49^. The CatBoost model moderately predict profile membership using pre-AMI data, and SHAP analysis offered crucial insights into the specific feature-categories driving each prediction. This approach is paramount for clinical translation, as it allows us to move beyond a “black box” model and provide clinicians with the rationale behind a patient’s risk stratification. By linking these profiles to specific genetic mechanisms and biological pathways ^50–53^, our study offers a mechanistic explanation for the observed clinical differences. The unique molecular signatures of cellular stress and genomic instability within the profiles provide a compelling biological basis for their poor prognosis and represent potential targets for future therapeutic interventions. These results suggest that while a patient’s comorbidity trajectory, as captured by our profiles, reflects vulnerability to acute stressors, its true clinical utility lies in informing specific care interventions based on underlying biological pathways, rather than solely on generic risk prediction.

### 4.1. Clinical Implications

The identified distinct multimorbidity profiles carry implications for clinical practice and personalised medicine. For participants in the SMO-CARD group, characterised by the most complex multimorbidity, advanced frailty, and the poorest survival outcomes, aggressive, multidisciplinary management strategies may be needed. This may involve coordinated care among cardiologists, endocrinologists, geriatricians, and other specialists, focusing on comprehensive geriatric assessment and proactive care planning to address their complex needs and high healthcare utilisation. Conversely, the ACUTE-CARD and CARDIOMIX profiles can benefit from targeted interventions focusing on cardiovascular-metabolic control and management of multisystemic issues, respectively. By stratifying AMI patients into these data-driven profiles, clinicians can tailor preventive measures, monitor high-risk individuals more closely, and potentially improve patient outcomes through truly personalised care plans. This stratification also has the potential to inform resource allocation within healthcare systems, ensuring that patients with the most complex needs receive appropriate levels of care. However, successful practical implementation will require further validation to seamlessly integrate these profiles into existing clinical workflows.

### 4.2. Limitations

Despite its strengths, this study has several limitations. First, the reliance on UK Biobank data, which primarily includes White British participants from the United Kingdom, may limit the generalisability of these findings to more diverse global populations with different demographic characteristics or healthcare contexts. Second, our focus on ICD codes, while comprehensive for diagnoses, may not capture the full spectrum of a participant’s health status, such as detailed disease severity, specific lifestyle factors, or undocumented conditions. Third, the retrospective nature of the data means that causal relationships cannot be definitively established, only associations. Furthermore, while our analysis links multimorbidity profiles to genetic and biological pathways, this connection is associative rather than causal. Future research would require dedicated experimental studies to validate the mechanistic hypotheses we have generated. Despite these limitations, our study provides a foundational framework that can be adapted and expanded upon, offering a significant step forward in understanding the complexities of multimorbidity in AMI patients.

### 4.3. Future Directions

Future research should prioritise validating these multimorbidity profiles in diverse populations to assess their generalisability across different demographic and healthcare settings. Integrating additional data types, such as genomic, metabolomic, proteomic, and more granular lifestyle data, could further refine the identified profiles and provide a more holistic understanding of multimorbidity. For instance, incorporating specific genetic markers, or polygenetic risk scores could reveal underlying biological predispositions that contribute to the development of multimorbidity profiles. Longitudinal studies are also essential to evaluate the real-world impact of profile-specific interventions on patient outcomes, such as whether tailored treatments for SMO-CARD participants can significantly improve their survival rates or reduce healthcare utilisation. Ultimately, exploring the integration of these profiles into clinical decision support systems could facilitate their real-time application in healthcare settings, potentially transforming how clinicians manage multimorbid AMI patients and paving the way for true precision medicine in cardiovascular care.

## 5. Conclusion

This study provides a comprehensive and interpretable framework for understanding multimorbidity in AMI patients. By leveraging temporal clustering, we have demonstrated that a participant’s disease trajectory, not just their static risk factors, defines clinically distinct subgroups with unique prognostic and biological signatures. Our findings highlight a critical clinical insight: traditional risk scores alone are not enough to capture the acute, multisystemic nature of AMI patients. We have shown that multimorbidity profiles when combined with established risk scores can enhance risk predictions in AMI participants. The transparency offered by our explainable AI models provides a clear rationale for these risk stratifications, which is essential for clinical adoption. Ultimately, this work represents a significant step toward a new paradigm in precision medicine for AMI, where personalised management strategies are informed by a deeper understanding of a patient’s dynamic multimorbidity profile.

## Data Availability Statement

This study was conducted using data from the UK Biobank under approved Application Number 83988. The UK Biobank dataset is not publicly available due to participant privacy protections and data governance restrictions. Researchers may apply for access to the UK Biobank resource through the established application process at: https://www.ukbiobank.ac.uk/enable-your-research/apply-for-access

Derived data products (including cluster labels, aggregated feature tables, and model outputs) generated during this study may be shared upon reasonable request to the corresponding author, subject to UK Biobank’s data sharing policies and ethical approval requirements. No individual-level data can be shared.

---

**Supplementary Figure 1:**
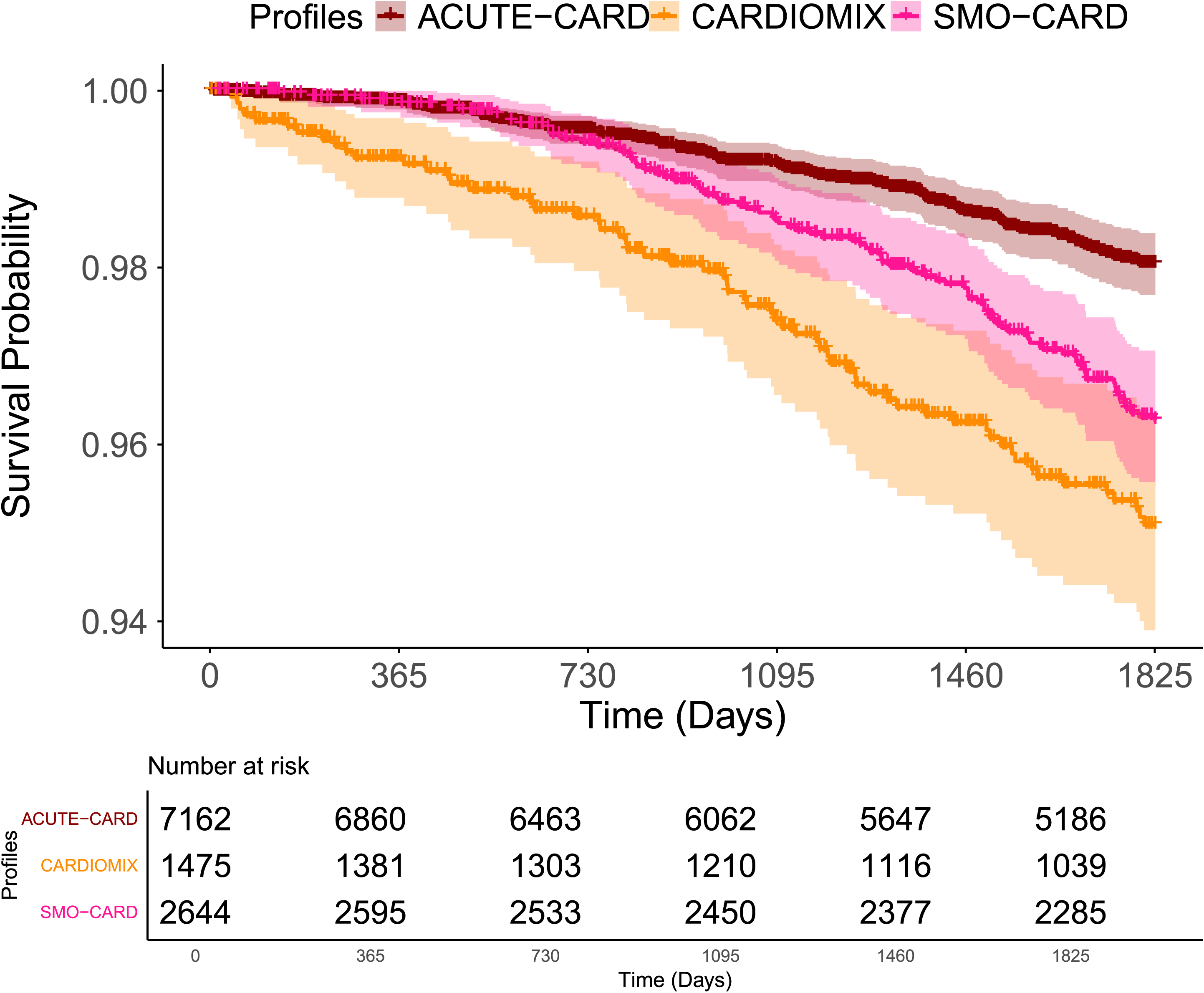
Kaplan-Meier curves for composite events, stratified by multimorbidity profiles. The Kaplan-Meier of the profiles were considered for All-cause events (within five years). The survival curves are colour-coded for each profile: ACUTE-CARD (dark red), CARDIOMIX (orange), and SMO-CARD (pink).

**Supplementary Figure 2:**
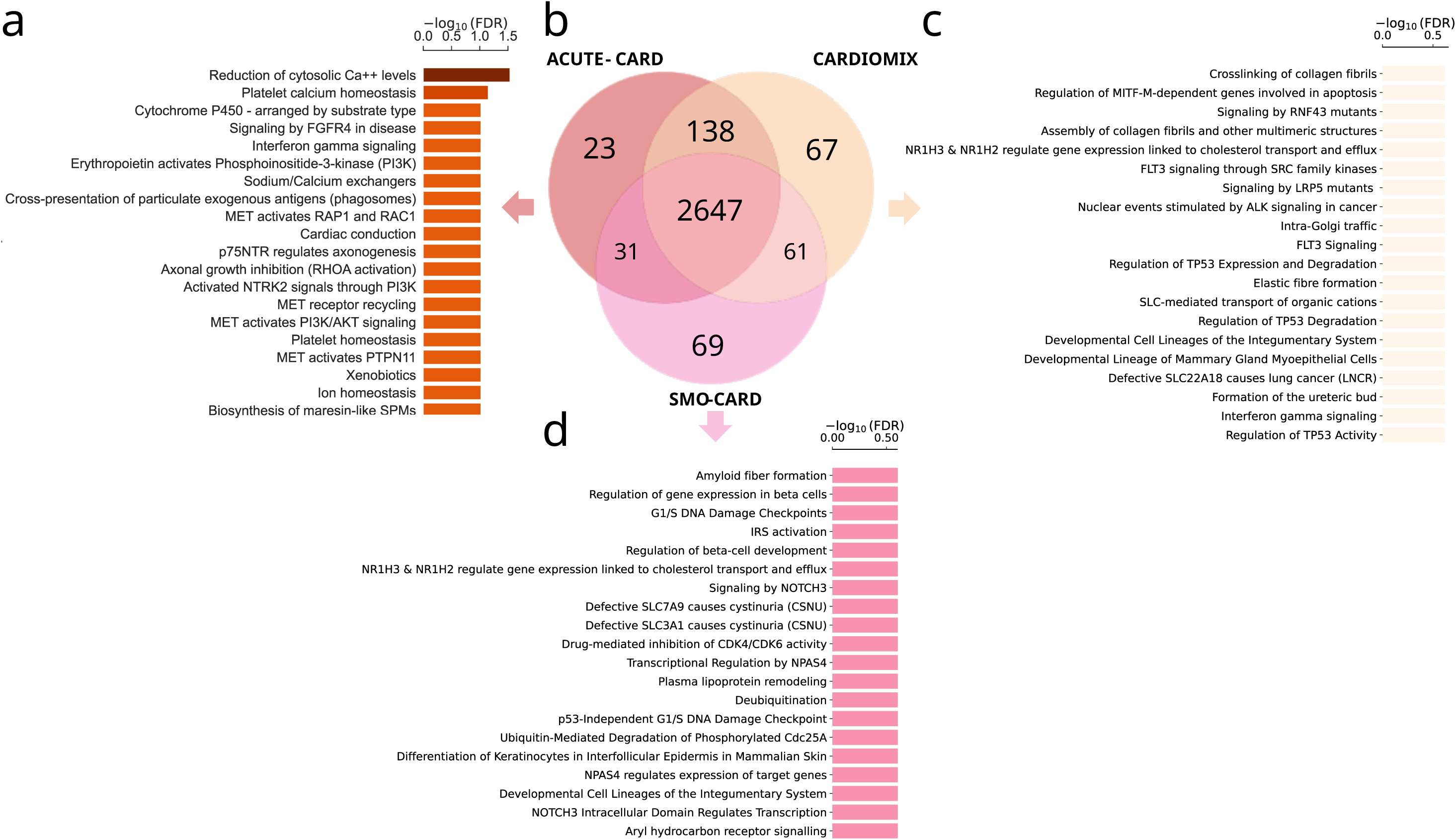
Comparative Pathway Enrichment and Gene Overlap Analysis of AMI -Associated SNPs Across ACUTE -CARD, CARDIOMIX, and SMO-CARD. Multi-panel figure illustrating gene overlap and pathway enrichment for genes containing acute AMI-associated SNPs in three profiles. (a) Top enriched Reactome pathways specific to the ACUTE-CARD, ranked by -log₁₀(FDR) (dark red to orange bars). (b) Venn diagram showing counts of overlapping genes across ACUTE-CARD, CARDIOMIX, and SMO-CARD profiles. (c) Top enriched Reactome pathways specific to the CARDIOMIX, ranked by -log₁₀(FDR) (yellow-orange bars). (d) Top enriched Reactome pathways specific to the SMO-CARD cohort or a complementary gene set, ranked by -log₁₀(FDR) (pink bars).

**Supplementary Figure 3:**
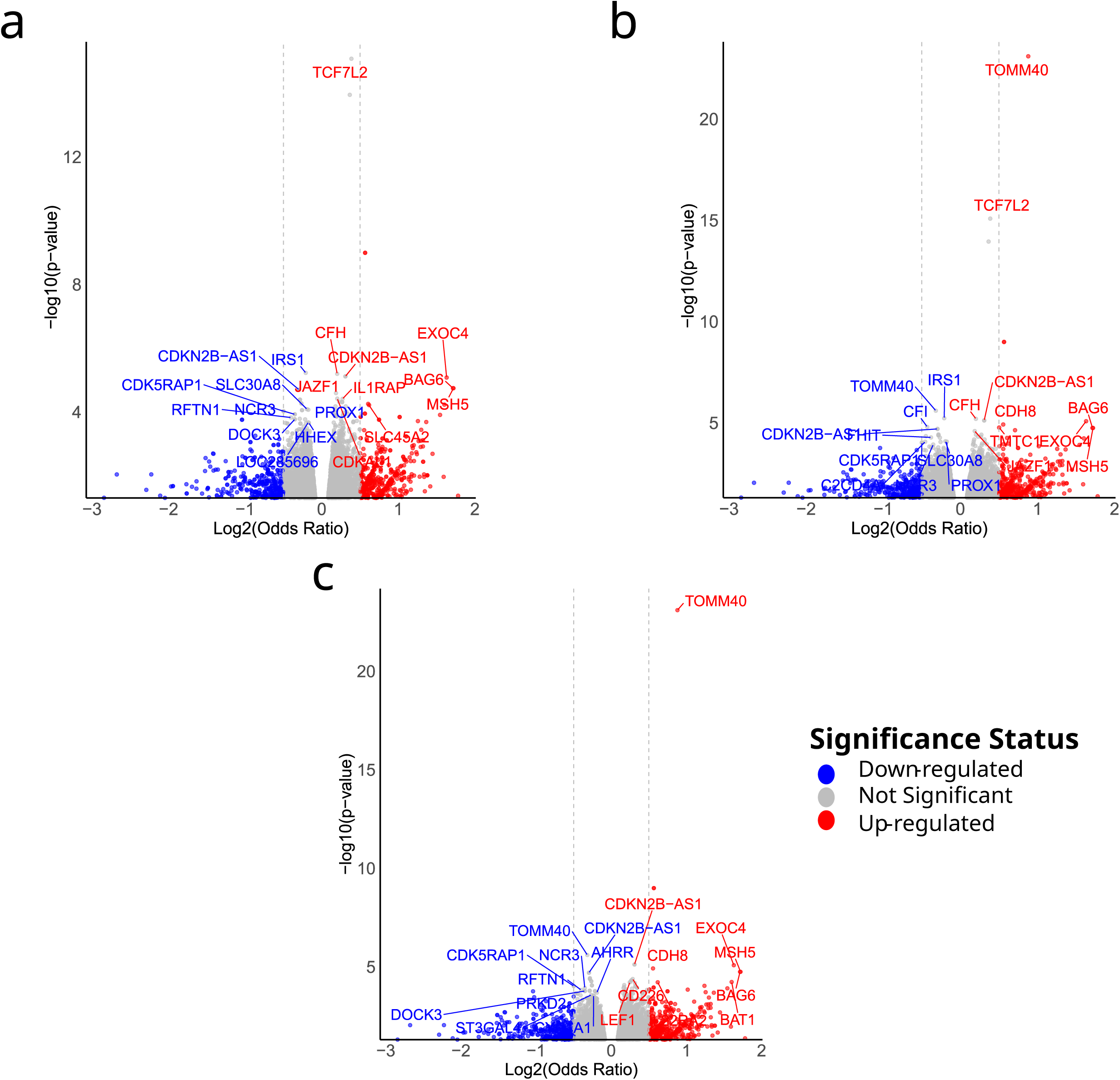
Statistical significance and effect size of genetic associations from a PheWAS analysis across the profiles: (a) ACUTE-CARD, (b) CARDIOMIX, and (c) SMO-CARD. X-axis: effect size (log₂OR), Y-axis: significance (−log₁₀p). Dashed lines mark thresholds: log₂OR=±0.5 (vertical) and p=0.05 (horizontal). Red = significant positive associations, blue = significant negative associations, grey = non-significant. Labels show top 10 most significant up and down-regulated genes.

## References

1. Boersma, E. et al. Acute myocardial infarction. The Lancet 361, 847–858 (2003).

2. Reed, G. W., Rossi, J. E. & Cannon, C. P. Acute myocardial infarction. The Lancet 389, 197–210 (2017).

3. Thygesen, K., Alpert, J. S. & White, H. D. Universal Definition of Myocardial Infarction. J Am Coll Cardiol 50, 2173–2195 (2007).

4. Roth, G. A. et al. Global Burden of Cardiovascular Diseases and Risk Factors, 1990–2019: Update From the GBD 2019 Study. J Am Coll Cardiol 76, 2982–3021 (2020).

5. Grey, C. et al. One in four major ischaemic heart disease events are fatal and 60% are pre-hospital deaths: a national data-linkage study (ANZACS-QI 8). Eur Heart J 38, 172–180 (2017).

6. Kochar, A. et al. Long-term mortality of older patients with acute myocardial infarction treated in US clinical practice. J Am Heart Assoc 7, (2018).

7. Coronary heart disease (Ischaemic heart disease) - types, causes & symptoms - BHF. https://www.bhf.org.uk/informationsupport/conditions/coronary-heart-disease.

8. Abdul Jabbar, A. Bin, Klisares, M., Gilkeson, K. & Aboeata, A. Acute Myocardial Infarction Mortality in the Older Population of the United States: An Analysis of Demographic and Regional Trends and Disparities from 1999 to 2022. J Clin Med 14, (2025).

9. Li, H. et al. Demographic and regional trends of acute myocardial infarction-related mortality among young adults in the US, 1999–2020. npj Cardiovascular Health 2025 2:*1* 2, 9- (2025).

10. Zuin, M. et al. Trends in acute myocardial infarction mortality in the European Union, 2012–2020. Eur J Prev Cardiol 30, 1758–1771 (2023).

11. Rahimi, K., Duncan, M., Pitcher, A., Emdin, C. A. & Goldacre, M. J. Mortality from heart failure, acute myocardial infarction and other ischaemic heart disease in England and Oxford: a trend study of multiple-cause-coded death certification. J Epidemiol Community Health 69, 1000–1005 (2015).

12. Shu, T. et al. Assessing Global, Regional, and National Time Trends and Associated Risk Factors of the Mortality in Ischemic Heart Disease Through Global Burden of Disease 2019 Study: Population-Based Study. JMIR Public Health Surveill 10, e46821 (2024).

13. Wang, W. et al. Global Burden of Disease Study 2019 suggests that metabolic risk factors are the leading drivers of the burden of ischemic heart disease. Cell Metab 33, 1943–1956.e2 (2021).

14. Smilowitz, N. R., Banco, D., Katz, S. D., Beckman, J. A. & Berger, J. S. Association between heart failure and perioperative outcomes in patients undergoing non-cardiac surgery. Eur Heart J Qual Care Clin Outcomes 7, 68–75 (2021).

15. Szummer, K. et al. Improved outcomes in patients with ST-elevation myocardial infarction during the last 20 years are related to implementation of evidence-based treatments: experiences from the SWEDEHEART registry 1995–2014. Eur Heart J 38, 3056–3065 (2017).

16. Zheng, W. et al. Impact of multimorbidity patterns on outcomes and treatment in patients with coronary artery disease. European Heart Journal Open 4, (2024).

17. Hartikainen, T. et al. Acute Myocardial Infarction. Euro Heart J 41, 2209–2216 (2023).

18. Wang, H., Zhao, T., Wei, X., Lu, H. & Lin, X. The prevalence of 30-day readmission after acute myocardial infarction: A systematic review and meta-analysis. Clin Cardiol 42, 889–898 (2019).

19. Bagai, A. et al. Multimorbidity, functional impairment, and mortality in older patients stable after prior acute myocardial infarction: Insights from the TIGRIS registry. Clin Cardiol 45, 1277–1286 (2022).

20. Wei, M. Y., Leis, A. M., Vasilyev, A. & Kang, A. J. Development and validation of new multimorbidity-weighted index for ICD-10-coded electronic health record and claims data: an observational study. BMJ Open 14, (2024).

21. Hall, M. et al. Multimorbidity and survival for patients with acute myocardial infarction in England and Wales: Latent class analysis of a nationwide population-based cohort. PLoS Med 15, e1002501 (2018).

22. Simard, M., Rahme, E., Calfat, A. C. & Sirois, C. Multimorbidity measures from health administrative data using ICD system codes: A systematic review. Pharmacoepidemiol Drug Saf 31, 1–12 (2022).

23. Than, M. P. et al. Machine Learning to Predict the Likelihood of Acute Myocardial Infarction. Circulation 140, 899–909 (2019).

24. Chen, Z. et al. Prediction of Myocardial Infarction From Patient Features With Machine Learning. Front Cardiovasc Med 9, 754609 (2022).

25. Doudesis, D. et al. Machine learning for diagnosis of myocardial infarction using cardiac troponin concentrations. Nat Med 29, 1201 (2023).

26. Zhang, Q. et al. Signaling pathways and targeted therapy for myocardial infarction. Signal Transduct Target Ther 7, (2022).

27. Ding, L. et al. Data-driven clustering approach to identify novel phenotypes using multiple biomarkers in acute ischaemic stroke: A retrospective, multicentre cohort study. EClinicalMedicine 53, 101639 (2022).

28. Zador, Z., Landry, A., Cusimano, M. D. & Geifman, N. Multimorbidity states associated with higher mortality rates in organ dysfunction and sepsis: A data-driven analysis in critical care. Crit Care 23, (2019).

29. Haue, A. D. et al. Subgrouping multimorbid patients with ischemic heart disease by means of unsupervised clustering: A cohort study of 72,249 patients defined comprehensively by diagnoses prior to presentation. medRxiv 2023.03.31.23288006 (2023) doi:10.1101/2023.03.31.23288006.

30. Delord, M. & Douiri, A. Unsupervised Clustering Approach to Multiple Time-to-Event Electronic Health Records applied to Multimorbidity associated with Myocardial Infarction. https://doi.org/10.21203/RS.3.RS-3127943/V1 (2023) doi:10.21203/RS.3.RS-3127943/V1.

31. Ibrahim, L., Mesinovic, M., Yang, K. W. & Eid, M. A. Explainable Prediction of Acute Myocardial Infarction Using Machine Learning and Shapley Values. IEEE Access 8, 210410–210417 (2020).

32. Wang, W., Lyu, G., Shi, Y. & Liang, X. Time Series Clustering Based on Dynamic Time Warping. Proceedings of the IEEE International Conference on Software Engineering and Service Sciences, ICSESS 2018-November, 487–490 (2018).

33. UK Biobank - UK Biobank. https://www.ukbiobank.ac.uk/.

34. Category 76. https://biobank.ndph.ox.ac.uk/showcase/label.cgi?id=76.

35. International Classification of Diseases (ICD) Information Sheet.

36. Vamathevan, J. et al. Applications of machine learning in drug discovery and development. Nat Rev Drug Discov 18, 463–477 (2019).

37. Rácz, A., Bajusz, D. & Héberger, K. Multi-Level Comparison of Machine Learning Classifiers and Their Performance Metrics. Molecules 2019, *Vol.* 24, *Page* 2811 24, 2811 (2019).

38. Wojtuch, A., Jankowski, R. & Podlewska, S. How can SHAP values help to shape metabolic stability of chemical compounds? J Cheminform 13, 1–20 (2021).

39. Kaasenbrood, L. et al. Distribution of Estimated 10-Year Risk of Recurrent Vascular Events and Residual Risk in a Secondary Prevention Population. Circulation 134, 1419–1429 (2016).

40. PheWAS Resources. https://phewascatalog.org/phewas/#home.

41. Hebbring, S. J. et al. A PheWAS approach in studying HLA-DRB1*1501. Genes & Immunity 2013 14:*3* 14, 187–191 (2013).

42. Jassal, B. et al. The reactome pathway knowledgebase. Nucleic Acids Res 48, D498–D503 (2020).

43. Reactome | Pathway Browser. https://reactome.org/PathwayBrowser/.

44. Moore, A. & Bell, M. XGBoost, A Novel Explainable AI Technique, in the Prediction of Myocardial Infarction: A UK Biobank Cohort Study. Clin Med Insights Cardiol 16, (2022).

45. Holzinger, A., Langs, G., Denk, H., Zatloukal, K. & Müller, H. Causability and explainability of artificial intelligence in medicine. Wiley Interdiscip Rev Data Min Knowl Discov 9, e1312 (2019).

46. Slack, D., Hilgard, S., Singh, S. & Lakkaraju, H. Reliable Post hoc Explanations: Modeling Uncertainty in Explainability. Adv Neural Inf Process Syst 34, 9391–9404 (2021).

47. Rawal, A., Raglin, A. J., Sadler, B. M. & Rawat, D. B. Explainability and causality for robust, fair, and trustworthy artificial reasoning. 10.1117/12.2666085 12538, 493–500 (2023).

48. Islam, M. A., Nittala, K. & Bajwa, G. Adding Explainability to Machine Learning Models to Detect Chronic Kidney Disease. Proceedings - 2022 IEEE 23rd International Conference on Information Reuse and Integration for Data Science, IRI 2022 297–302 (2022) doi:10.1109/IRI54793.2022.00069.

49. London, A. J. Artificial Intelligence and Black-Box Medical Decisions: Accuracy versus Explainability. Hastings Center Report 49, 15–21 (2019).

50. Feng, Q. et al. The role of major immune cells in myocardial infarction. Front Immunol 13, 1084460 (2023).

51. Matter, M. A. et al. Inflammation in acute myocardial infarction: the good, the bad and the ugly. Eur Heart J 45, 89–103 (2024).

52. Luo, J. et al. Gene regulatory network analysis identifies key genes and regulatory mechanisms involved in acute myocardial infarction using bulk and single cell RNA-seq data. bioRxiv 2021.08.26.457775 (2021) doi:10.1101/2021.08.26.457775.

53. Ruparelia, N. et al. Acute myocardial infarction activates distinct inflammation and proliferation pathways in circulating monocytes, prior to recruitment, and identified through conserved transcriptional responses in mice and humans. Eur Heart J 36, 1923–1934 (2015).

