## Supplementary Table 1 - 5 for "Leveraging Explainable Temporal-Modelling Machine Learning to Identify Distinct Multimorbidity Trajectory Profiles in Acute Myocardial Infarction"

**Supplementary Table 1:** Association of Post-AMI Disease-specific Clusters With ICD-10 Themes Derived from Latent Dirichlet Allocation (LDA) Analysis

| **ICD10 code** | **Health condition** | **Theme** | **ACUTE-CARD (count)** | **CARDIOMIX (count)** | **SMO-CARD (count)** | **Total count** | **p-value** | **FDR adj p-value** |
| --- | --- | --- | --- | --- | --- | --- | --- | --- |
| I25 | Chronic ischemic heart disease | Theme 1, Theme 2 | 3472 | 264 | 5250 | 8986 | <0.0001 | <0.0001 |
| Z95 | Presence of cardiac and vascular implants and grafts | Theme 1, Theme 2 | 2534 | 31 | 2258 | 4823 | <0.0001 | <0.0001 |
| I10 | Essential (primary) hypertension | Theme 1, Theme 2 | 1578 | 19 | 2174 | 3771 | <0.0001 | <0.0001 |
| E78 | Disorders of lipoprotein metabolism and other lipidemias | Theme 1, Theme 2 | 1541 | 19 | 2202 | 3762 | <0.0001 | <0.0001 |
| Z82 | Family history of certain disabilities and chronic diseases (leading to disablement) | Theme 1, Theme 2 | 1116 | 24 | 1982 | 3122 | <0.0001 | <0.0001 |
| I20 | Angina pectoris | Theme 1, Theme 2 | 1364 | 11 | 1380 | 2755 | <0.0001 | <0.0001 |
| I51 | Complications and ill-defined descriptions of heart disease | Theme 1, Theme 2 | 1528 | 25 | 1061 | 2614 | <0.0001 | <0.0001 |
| I50 | Heart failure | Theme 1, Theme 2 | 1657 | 27 | 803 | 2487 | <0.0001 | <0.0001 |
| R07 | Pain in throat and chest | Theme 1, Theme 2 | 1093 | 7 | 840 | 1940 | <0.0001 | <0.0001 |
| I48 | Atrial fibrillation and flutter | Theme 1, Theme 2 | 1250 | 15 | 535 | 1800 | <0.0001 | <0.0001 |
| I24 | Other acute ischemic heart diseases | Theme 1, Theme 2 | 1006 | 12 | 709 | 1727 | <0.0001 | <0.0001 |
| N17 | Acute kidney failure | Theme 1, Theme 2 | 1283 | 8 | 151 | 1442 | <0.0001 | <0.0001 |
| E87 | Other disorders of fluid, electrolyte and acid-base balance | Theme 1, Theme 2 | 1156 | 7 | 201 | 1364 | <0.0001 | <0.0001 |
| I08 | Multiple valve diseases | Theme 1, Theme 2 | 841 | 7 | 510 | 1358 | <0.0001 | <0.0001 |
| I95 | Hypotension | Theme 1, Theme 2 | 1044 | 7 | 276 | 1327 | <0.0001 | <0.0001 |
| E66 | Overweight and obesity | Theme 1, Theme 2 | 719 | 9 | 542 | 1270 | <0.0001 | <0.0001 |
| I44 | Atrioventricular and left bundle-branch block | Theme 1, Theme 2 | 774 | 11 | 385 | 1170 | <0.0001 | <0.0001 |
| J18 | Pneumonia, unspecified organism | Theme 1, Theme 2 | 988 | 12 | 150 | 1150 | <0.0001 | <0.0001 |
| R00 | Abnormalities of heartbeat | Theme 1, Theme 2 | 697 | 7 | 383 | 1087 | <0.0001 | <0.0001 |
| F17 | Nicotine dependence | Theme 2 | 450 | 6 | 612 | 1068 | <0.0001 | <0.0001 |
| N18 | Chronic kidney disease (CKD) | Theme 1, Theme 2 | 808 | 7 | 217 | 1032 | <0.0001 | <0.0001 |
| Y83 | Surgical operation and other surgical procedures as the cause of abnormal reaction of the patient, or of later complication, without mention of misadventure at the time of the procedure | Theme 1, Theme 2 | 808 | 5 | 182 | 995 | <0.0001 | <0.0001 |
| D64 | Other anemias | Theme 1, Theme 2 | 862 | 4 | 129 | 995 | <0.0001 | <0.0001 |
| K21 | Gastro-esophageal reflux disease | Theme 1, Theme 2 | 629 | 3 | 361 | 993 | <0.0001 | <0.0001 |
| J90 | Pleural effusion, not elsewhere classified | Theme 1, Theme 2 | 848 | 8 | 119 | 975 | <0.0001 | <0.0001 |

Acute Cardiorenal–Respiratory with Chronic Metabolic Disease (ACUTE-CARD), Cardiometabolic with Mixed Arrhythmic–Ischaemic Burden (CARDIOMIX), Smoking-Related Cardiovascular Disease with Multimorbidity (SMO-CARD). p-value: The statistical significance of the difference in prevalence of the health condition across the three profiles. FDR adj p-value: The p-value adjusted for multiple testing using the False Discovery Rate (FDR) method. This adjustment helps to reduce the chance of false positives. Themes were derived from an LDA topic-modelling analysis of ICD-10 codes to identify clinically coherent clusters of comorbidities.

**Supplementary Table 2:** Weighted Average scores of performance metrics in the testing set

| **Classifier** | **Accuracy** | **Precision** | **Recall** | **F1-score** | **AUROC** |
| --- | --- | --- | --- | --- | --- |
| Logistic Regression | 0.62 | 0.68 | 0.62 | 0.61 | 0.76 |
| Random Forest | 0.67 | 0.64 | 0.67 | 0.65 | 0.76 |
| XGBoost | 0.63 | 0.67 | 0.63 | 0.64 | 0.77 |
| CatBoost | 0.65 | 0.67 | 0.65 | 0.65 | 0.77 |

Note: Weighted average scores were used to evaluate the multiclass performance.

**Supplementary Table 3:** Key Health Conditions Across Multimorbidity Profiles in pre-AMI data

| **ICD10 code** | **Health condition** | **ACUTE-CARD (count)** | **CARDIOMIX (count)** | **SMO-CARD (count)** | **ACUTE-CARD prevalence** | **CARDIOMIX prevalence** | **SMO-CARD prevalence** | **Total count** | **p-value** | **FDR adj**  **p-value** |
| --- | --- | --- | --- | --- | --- | --- | --- | --- | --- | --- |
| I25 | Chronic ischemic heart disease | 2371 | 277 | 4037 | 0.45 | 0.30 | 0.62 | 6685 | <0.0001 | <0.0001 |
| E78 | Disorders of lipoprotein metabolism and other lipidemias | 1037 | 54 | 1803 | 0.2 | 0.06 | 0.28 | 2894 | <0.0001 | <0.0001 |
| I10 | Essential (primary) hypertension | 1001 | 67 | 1721 | 0.19 | 0.07 | 0.27 | 2789 | <0.0001 | <0.0001 |
| Z82 | Family history of certain disabilities and chronic diseases (leading to disablement) | 838 | 52 | 1710 | 0.16 | 0.06 | 0.26 | 2600 | <0.0001 | <0.0001 |
| I51 | Complications and ill-defined descriptions of heart disease | 884 | 58 | 855 | 0.17 | 0.06 | 0.13 | 1797 | <0.0001 | <0.0001 |
| I50 | Heart failure | 934 | 60 | 650 | 0.18 | 0.07 | 0.1 | 1644 | <0.0001 | <0.0001 |
| I20 | Angina pectoris | 628 | 47 | 706 | 0.12 | 0.05 | 0.11 | 1381 | <0.0001 | <0.0001 |
| I48 | Atrial fibrillation and flutter | 671 | 51 | 399 | 0.13 | 0.06 | 0.06 | 1121 | <0.0001 | <0.0001 |
| Z95 | Presence of cardiac and vascular implants and grafts | 436 | 49 | 571 | 0.08 | 0.05 | 0.09 | 1056 | 0.0015 | 0.0078 |
| F17 | Nicotine dependence | 405 | 18 | 601 | 0.08 | 0.02 | 0.09 | 1024 | <0.0001 | <0.0001 |
| I08 | Multiple valve diseases | 504 | 17 | 423 | 0.1 | 0.02 | 0.07 | 944 | <0.0001 | <0.0001 |
| I44 | Atrioventricular and left bundle-branch block | 474 | 29 | 342 | 0.09 | 0.03 | 0.05 | 845 | <0.0001 | <0.0001 |
| N17 | Acute kidney failure | 596 | 38 | 170 | 0.11 | 0.04 | 0.03 | 804 | <0.0001 | <0.0001 |
| E11 | Type 2 diabetes mellitus | 373 | 32 | 378 | 0.07 | 0.03 | 0.06 | 783 | <0.0001 | 0.0003 |
| R07 | Pain in throat and chest | 368 | 47 | 366 | 0.07 | 0.05 | 0.06 | 781 | 0.005 | 0.0204 |
| E66 | Overweight and obesity | 349 | 22 | 402 | 0.07 | 0.02 | 0.06 | 773 | <0.0001 | <0.0001 |
| I24 | Other acute ischemic heart diseases | 356 | 45 | 352 | 0.07 | 0.05 | 0.05 | 753 | 0.0045 | 0.0187 |
| E87 | Other disorders of fluid, electrolyte and acid-base balance | 502 | 31 | 201 | 0.09 | 0.03 | 0.03 | 734 | <0.0001 | <0.0001 |
| J18 | Pneumonia, unspecified organism | 499 | 47 | 139 | 0.09 | 0.05 | 0.02 | 685 | <0.0001 | <0.0001 |
| I46 | Cardiac arrest | 274 | 39 | 326 | 0.05 | 0.04 | 0.05 | 639 | 0.4779 | 0.6429 |

Acute Cardiorenal–Respiratory with Chronic Metabolic Disease (ACUTE-CARD), Cardiometabolic with Mixed Arrhythmic–Ischaemic Burden (CARDIOMIX), Smoking-Related Cardiovascular Disease with Multimorbidity (SMO-CARD). Disease prevalence (%): The percentage of participants within each profile who have the specified health condition, p-value: The statistical significance of the difference in prevalence of the health condition across the three profiles. FDR adj p-value: The p-value adjusted for multiple testing using the False Discovery Rate (FDR) method. This adjustment helps to reduce the chance of false positives.

**Supplementary Table 4:** Key Health Conditions Across Multimorbidity Profiles in post-AMI data

| **ICD10 code** | **Health condition** | **ACUTE-CARD (count)** | **CARDIOMIX (count)** | **SMO-CARD (count)** | **ACUTE-CARD prevalence** | **CARDIOMIX prevalence** | **SMO-CARD prevalence** | **Total count** | **p-value** | **FDR adj p-value** |
| --- | --- | --- | --- | --- | --- | --- | --- | --- | --- | --- |
| I25 | Chronic ischemic heart disease | 3472 | 264 | 5250 | 0.66 | 0.29 | 0.81 | 8986 | <0.0001 | <0.0001 |
| Z95 | Presence of cardiac and vascular implants and grafts | 2534 | 31 | 2258 | 0.48 | 0.03 | 0.35 | 4823 | <0.0001 | <0.0001 |
| I10 | Essential (primary) hypertension | 1578 | 19 | 2174 | 0.3 | 0.02 | 0.34 | 3771 | <0.0001 | <0.0001 |
| E78 | Disorders of lipoprotein metabolism and other lipidemias | 1541 | 19 | 2202 | 0.29 | 0.02 | 0.34 | 3762 | <0.0001 | <0.0001 |
| Z82 | Family history of certain disabilities and chronic diseases (leading to disablement) | 1116 | 24 | 1982 | 0.21 | 0.03 | 0.31 | 3122 | <0.0001 | <0.0001 |
| I20 | Angina pectoris | 1364 | 11 | 1380 | 0.26 | 0.01 | 0.21 | 2755 | <0.0001 | <0.0001 |
| I51 | Complications and ill-defined descriptions of heart disease | 1528 | 25 | 1061 | 0.29 | 0.03 | 0.16 | 2614 | <0.0001 | <0.0001 |
| I50 | Heart failure | 1657 | 27 | 803 | 0.31 | 0.03 | 0.12 | 2487 | <0.0001 | <0.0001 |
| R07 | Pain in throat and chest | 1093 | 7 | 840 | 0.21 | 0.01 | 0.13 | 1940 | <0.0001 | <0.0001 |
| I48 | Atrial fibrillation and flutter | 1250 | 15 | 535 | 0.24 | 0.02 | 0.08 | 1800 | <0.0001 | <0.0001 |
| I24 | Other acute ischemic heart diseases | 1006 | 12 | 709 | 0.19 | 0.01 | 0.11 | 1727 | <0.0001 | <0.0001 |
| N17 | Acute kidney failure | 1283 | 8 | 151 | 0.24 | 0.01 | 0.02 | 1442 | <0.0001 | <0.0001 |
| E87 | Other disorders of fluid, electrolyte and acid-base balance | 1156 | 7 | 201 | 0.22 | 0.01 | 0.03 | 1364 | <0.0001 | <0.0001 |
| I08 | Multiple valve diseases | 841 | 7 | 510 | 0.16 | 0.01 | 0.08 | 1358 | <0.0001 | <0.0001 |
| I95 | Hypotension | 1044 | 7 | 276 | 0.2 | 0.01 | 0.04 | 1327 | <0.0001 | <0.0001 |
| E66 | Overweight and obesity | 719 | 9 | 542 | 0.14 | 0.01 | 0.08 | 1270 | <0.0001 | <0.0001 |
| I44 | Atrioventricular and left bundle-branch block | 774 | 11 | 385 | 0.15 | 0.01 | 0.06 | 1170 | <0.0001 | <0.0001 |
| J18 | Pneumonia, unspecified organism | 988 | 12 | 150 | 0.19 | 0.01 | 0.02 | 1150 | <0.0001 | <0.0001 |
| R00 | Abnormalities of heartbeat | 697 | 7 | 383 | 0.13 | 0.01 | 0.06 | 1087 | <0.0001 | <0.0001 |
| F17 | Nicotine dependence | 450 | 6 | 612 | 0.09 | 0.01 | 0.09 | 1068 | <0.0001 | <0.0001 |

Acute Cardiorenal–Respiratory with Chronic Metabolic Disease (ACUTE-CARD), Cardiometabolic with Mixed Arrhythmic–Ischaemic Burden (CARDIOMIX), Smoking-Related Cardiovascular Disease with Multimorbidity (SMO-CARD). Disease prevalence (%): The percentage of participants within each profile who have the specified health condition, p-value: The statistical significance of the difference in prevalence of the health condition across the three profiles. FDR adj p-value: The p-value adjusted for multiple testing using the False Discovery Rate (FDR) method. This adjustment helps to reduce the chance of false positives.

**Supplementary Table 5**: Cox proportional hazards models (Models 1–4) for 0–5-year all-cause mortality

| **Model** | **Covariate** | **HR (95% CI)** | **p-value** |
| --- | --- | --- | --- |
| 1. Profiles only | Profile (per unit increase) | 1.40 (1.23 – 1.59) | <0.001 |
| 2. SMART-risk only | SMART-risk score (per unit) | 1.05 (1.04 – 1.07) | <0.001 |
| 3. Profiles + SMART-risk | Profile (per unit increase) | 1.25 (1.10 – 1.43) | 0.001 |
|  | SMART-risk score (per unit) | 1.05 (1.04 – 1.06) | <0.001 |
| 4. Fully adjusted (Profile + SMART-risk + covariates) | Profile (per unit increase) | 1.12 (0.98 – 1.29) | 0.105 |
|  | SMART-risk score | 1.03 (1.02 – 1.05) | <0.001 |
|  | Age at baseline | 1.06 (1.03 – 1.08) | <0.001 |
|  | Sex (male) | 0.71 (0.54 – 0.95) | 0.019 |
|  | IMD score | 1.01 (1.00 – 1.02) | 0.002 |
|  | BMI | 1.00 (0.97 – 1.02) | 0.739 |
|  | Diabetes | 1.50 (1.04 – 2.17) | 0.031 |
|  | CAD | 1.21 (0.84 – 1.72) | 0.302 |
|  | Cerebrovascular disease | 1.06 (0.53 – 2.15) | 0.862 |
|  | PAD | 1.17 (0.61 – 2.23) | 0.638 |

Values are hazard ratios (HR) with 95% confidence intervals. Reference profile = ACUTE-CARD. Model D additionally adjusts for all listed covariates.
